# *LRRK2* p.G2385R and p.R1628P Variants in a Multi-Ethnic Asian Parkinson’s Cohort: Epidemiology and Clinical Insights

**DOI:** 10.1101/2025.08.06.25332985

**Authors:** Jun Wen Goh, Jia Lun Lim, Rui Yan Ong, Qing Hui Yong, Cindy Choey Yee Lew, Tzi Shin Toh, Jia Wei Hor, Yi Wen Tay, Jannah Zulkefli, Anis Nadhirah Khairul Anuar, Hans Xing Ding, Jie Ping Schee, Yuan Ye Beh, Khairul Azmi Ibrahim, Ahmad Shahir Mawardi, Thien Thien Lim, Irene Looi, Yuen Kang Chia, Joshua Chin Ern Ooi, Wan Chung Law, Siaw Cheng Wong, Yue Hui Lau, Pei Chiek Teh, Tien Lee Ong, Wee Kooi Cheah, Esther Sammler, Shalini Padmanabhan, Lei Cheng Lit, Eng King Tan, Azlina Ahmad-Annuar, Shen-Yang Lim, Ai Huey Tan

## Abstract

The frequency and clinical impact of *LRRK2* p.G2385R and p.R1628P risk variants in Parkinson’s disease (PD) remain uncertain, particularly across different Asian populations. We genotyped 3,058 multi-ethnic Malaysian PD patients, performed detailed phenotyping in 185, and analyzed disease progression in 635 using longitudinal Clinical Impression of Severity Index for PD scores. p.G2385R was largely confined to Chinese (8.2%), while p.R1628P occurred in mixed ancestry (11.0%), Chinese (8.3%), Malays (7.7%), and is reported for the first time in indigenous groups (3.9%). Double-variant carriers had younger onset and more frequently had positive family history. Compared with non-carriers, p.R1628P carriers had lower rates of dementia and orthostatic hypotension, and slower progression of motor signs and disability. Our findings highlight ethnic differences in the distribution of *LRRK2* Asian variants, and suggest that these variants influence onset age, familial occurrence, non-motor features, and disease course, with implications for personalized approaches to PD in Asian populations.

## INTRODUCTION

Parkinson’s disease (PD) affects ∼11.8 million people worldwide,^1^ with disease incidence projected to continue rising, in part due to population aging.^1–3^ Asia will have more than 60% of the world’s population aged at least 65 years by the 2030s, and this region is poised to account for the majority of PD patients worldwide.^3^ Along with aging, environmental, and lifestyle factors, genetics plays an important role in PD development and phenotypic expression (clinical features and progression) of the disease.^2–5^ Genome-wide association studies have implicated >90 genetic risk loci for PD.^6^ Heterozygous pathogenic variants in the *LRRK2* gene (e.g., p.G2019S) are causal for PD, implicated in monogenic and familial forms, but these have only been relatively rarely reported in Asian populations.^7–10^ The *LRRK2* Asian risk variants, p.G2385R and p.R1628P, have been identified as key risk factors for sporadic PD in various Asian populations,^3,11–15^ including the Malaysian population.^16^

The frequency of these *LRRK2* Asian risk variants among PD patients varies substantially across studies, with the frequency of p.G2385R reportedly ranging from 7.5% among Chinese patients in Singapore^17^ to 18.6% in a small study in eastern China,^18^ and p.R1628P ranging from 3.0%^18^ to 8.4%^19^ in eastern China and Singapore, respectively. Studies in Japan found frequencies of 10.4-11.5% for p.G2385R, with p.R1628P being absent,^12,20^ with a similar pattern seen in Korea.^14,21,22^ These studies were limited in sample size (the vast majority with only a few hundred to less than one thousand PD patients),^18^ and most focused on the Chinese and Japanese populations,^23,24^ with limited data available for other Asian ancestry groups, such as the Malays, Indians, and indigenous groups.^11,15,25–28^

There is also limited research, with conflicting findings, investigating the association of the *LRRK2* Asian variants with clinical phenotypes. While clinical phenotype of p.G2385R has been more well studied (with more than 10 studies to date),^21,29–37^ studies on p.R1628P remain scarce and are often limited by small sample size.^15,37–39^ Notably, few studies have compared the clinical manifestation between p.G2385R and p.R1628P carriers,^40,41^ and some grouped p.G2385R and p.R1628P into one *LRRK2* group,^18,42,43^ which may potentially obscure distinctive clinical characteristics associated with each variant. Evidence on disease progression is even more limited, with only four studies reporting inconsistent findings in patients with p.G2385R and p.R1628P variants.^37,44–46^ Thus, it currently remains uncertain whether carriers of either variant have a different clinical phenotype and progression, against each other and compared to non-carriers.

There is a pressing need for more data from underrepresented multi-ethnic populations^47^ to better understand the role of *LRRK2* p.R1628P and p.G2385R risk variants in Asian PD, which will help to facilitate the delivery of personalized precision medicine based on genetic profiles.^5,8,48^ Reliable data on the frequency and clinical manifestations of *LRRK2*-related PD are particularly crucial at this juncture, as the PD field moves towards more targeted selection or stratification of patients for biomarker studies,^49^ and trials of novel genetics-informed therapies.^5,8,9,50,51^ To address these gaps in knowledge, we determined the frequency of the *LRRK2* p.G2385R and p.R1628P risk variants in a large (n>3000) cohort of different Asian ancestries (Chinese, Malay, Indian, and indigenous groups) and conducted a longitudinal study (up to 4.5 years) to investigate the clinical profiles and progression of the variant carriers compared to non-carriers.

## METHODS

### Subject recruitment

Patients clinically diagnosed with PD were recruited via convenience sampling by neurologists specializing, or with an interest, in movement disorders at fifteen tertiary or quaternary medical centers across Malaysia: Universiti Malaya Medical Centre (UMMC), Universiti Malaya Specialist Centre (UMSC), Queen Elizabeth Hospital Kota Kinabalu, Sultanah Nur Zahirah Hospital Kuala Terengganu, Seberang Jaya Hospital, Kuala Lumpur Hospital, Taiping Hospital, Seremban Hospital, Sarawak General Hospital, Tengku Ampuan Rahimah Hospital Klang, Sri Aman Hospital, Raja Permaisuri Bainun Hospital Ipoh, Sungai Buloh Hospital, Sibu Hospital, and Bintulu Hospital, with the first site (UMMC) commencing in 2010. Patients with all other causes of parkinsonism, including Parkinson-plus syndromes, drug-induced parkinsonism, and vascular parkinsonism, were excluded. Other exclusion criteria were known monogenic forms of PD, related to *SNCA*, *LRRK2* (pathogenic variant p.R1441C)*, PRKN*, *PINK1*, and *PARK7*/*DJ-1*.^4,5,52–55^ Ethics approval was obtained from the Medical Research Ethics Committee at UMMC and the Ministry of Health Malaysia (No. 20191010-7917 and NMRR-19-3762-52429). Written informed consent was obtained from all participants.

### Sample collection, DNA extraction, and genomic analysis

Ten milliliters of peripheral blood samples were collected in EDTA tubes and preserved at -20°C until DNA extraction. Alternatively, saliva samples were collected using Oragene® DNA collection kits (DNA Genotek® Oragene® OG-500). DNA extractions were performed using the Favorgen Prep™ Blood Cell Genomic DNA Extraction Maxi Kit, or the Oragene prep IT L2P extraction kit (saliva).

Genotyping of *LRRK2* p.G2385R (NM_198578.4; c.7153G>A) and p.R1628P (NM_198578.4; c.4883G>C) variants was performed using TaqMan allelic discrimination assays (C 63498855_10 and C 63497592_10 respectively) using the StepOnePlus™ Real-Time PCR (RTPCR) Systems platform following the manufacturer’s instructions, and genotype calling with the StepOne™ software. Genotypes were validated in a random selection of 10 samples per assay, by PCR and Sanger sequencing (primers available upon request) to determine the error rate.

### Clinical assessments

Basic clinico-demographic information was obtained on recruited subjects across all centers (n=3058) (referred to as the “overall cohort”), including age, gender, self-reported ancestry, age at PD diagnosis, and the presence or absence of family history of PD.

For detailed clinical phenotyping, a subset of UMMC and UMSC patients (n=185) were recruited as part of an international Michael J. Fox Foundation (MJFF)-funded biobanking project to study the clinical and biomarker features of patients carrying the Asian *LRRK2* variants p.R1628P and p.G2385R (referred to as the “MJFF Asian *LRRK2* cohort”). Variant carriers and an age-matched *LRRK2* p.G2385R and p.R1628P variant-negative PD control group underwent assessments of disease severity using the Movement Disorder Society Unified PD Rating Scale (MDS-UPDRS), Hoehn and Yahr staging (H&Y), the Clinical Impression of Severity Index for PD (CISI-PD), and Montreal Cognitive Assessment (MoCA). Additionally, a structured checklist for the presence or absence of 30 motor and non-motor features during the course of PD was also utilized, as previously described^54,56^ based on the recommendations of the Movement Disorder Society Genetic Mutation Database (MDSGene; www.mdsgene.org), which collates English-language reports of patients with monogenic movement disorders.^9,57^ Because of links between *LRRK2* and gastrointestinal (GI) disturbances,^58,59^ GI-related questionnaires including the Patient Assessment of Constipation-Symptoms (PAC-SYM) and ROME IV constipation questionnaire were also administered. MDS-UPDRS parts III (motor examination, in the ON-medication state) and IV (motor complications), H&Y, and CISI-PD were evaluated by experienced movement disorder neurologists (SYL and AHT), with the remaining assessments administered by trained research assistants, between September 2021 to August 2023.

For clinical progression, a separate but overlapping subset of patients who were under active follow-up at UMMC and UMSC and had longitudinal CISI-PD data (evaluated by SYL and AHT from February 2019 onwards) were analyzed (n=635) (hereon referred to as the “longitudinal cohort”). The CISI-PD evaluations were repeated at intervals of at least 1.5 years to evaluate PD progression and are ongoing (with some patients having three repeat CISI-PD evaluations done over a follow-up period of 4.5 years or more). The CISI-PD progression rates were determined by calculating the difference between the latest and the baseline CISI-PD scores, then dividing this difference by the follow-up period in months. This was analyzed for the total CISI-PD score, as well for each of the four component domains (motor signs, disability, motor complications, and cognitive status). The progression rates were thus recorded as change in scores per month.

### Statistical analysis

Data were analyzed using SPSS for Macintosh Version 26.0 (SPSS, Inc., Chicago, IL, USA). Normality testing was conducted using the Kolmogorov-Smirnov test. Between-group differences in demographics, genotypes, clinical profile, and progression were analyzed using Chi-Square, Fisher’s exact test, independent t-test, or Mann-Whitney U test, as appropriate. Quade test (non-parametric ANCOVA) was used to control for age and baseline CISI-PD scores in evaluating between-group differences in CISI-PD progression scores. Statistical significance was set at p<0.05.

## RESULTS

### Clinico-demographic characteristics of the overall cohort

After excluding patients with known monogenic PD and selecting only index cases, a total of 3058 PD patients were included in the analysis. The mean age at recruitment was 65.4 ± 10.2 (range: 27-95) years, with the majority being male (57.9%). The mean age at PD diagnosis was 60.8 ± 11.2 (range 16-91) years, whereby 476 (15.6%) of the participants were diagnosed before the age of 50 (early-onset PD, EOPD), with a mean age at PD diagnosis of 42.5 ± 5.5 years. A total of 545 (17.8%) participants had a confirmed or possible (based on self-report) family history of PD.

The majority of the cohort was of Chinese ancestry, accounting for 1930 (63.1%) of the participants. This was followed by Malay (16.6%, 507/3058), Indian (13.4%, 410/3058), mixed ancestries (e.g., Chinese-Malay, Chinese-Indian, Chinese-Kadazan, Chinese-Dutch, Chinese-Pakistani, Malay-Turkish) (3.6%, 109/3058) and various indigenous groups (3.4%, 103/3058). The indigenous group included patients of Bajau, Bidayuh, Bisaya, Bugis, Dusun, Iban, Iranun, Jawa, Kadazan, Kedayan, Lundayoh, Melanau, Murut, Rawa, Rungus, Sinau, Sungai, Suluk, and Tidung ancestry.

### Frequency, age at diagnosis and family history

For *LRRK2* p.G2385R, the overall frequency was 5.5% (168/3058), consisting mostly of heterozygous carriers (165/168, 5.4%), and only three (0.1%) homozygous carriers. The vast majority of the heterozygous p.G2385R carriers were Chinese (93.9%, 155/165), followed by mixed-ancestry patients (4.8%, 8/165) (all of whom reported having Chinese ancestry), and a very small number of Malays (1.2%, 2/165) patients. All three homozygous carriers were Chinese. None of the Indian or indigenous patients were p.G2385R variant carriers.

The overall frequency of the *LRRK2* p.R1628P was 7.1% (217/3058), again consisting mostly of heterozygous carriers (213/217, 7.0%), and only four (0.1%) homozygous carriers. The majority of heterozygous p.R1628P carriers were Chinese (75.6%, 158/213), followed by Malay (17.8%, 38/213), mixed ancestry (5.6%, 12/213), and indigenous groups (1.9%, 4/213: n=1/16 Bajau, n=1/7 Bugis, n=1/5 Sungai, n=1/28 Dusun), with one (0.5%) of Indian descent. Among the homozygous carriers, three were Chinese and one was Malay. Only 14 patients (0.5%) were double heterozygous for both p.R1628P and p.G2385R (n=13 Chinese, n=1 Malay-Chinese mixed ancestry).

Among the ancestral groups (Figure 1), the frequency of p.G2385R was highest among Chinese (8.2%, 158/1930), followed by the mixed-ancestry group (7.3%, 8/109) (all of whom reported having some Chinese ancestry). This variant was rare in the other ancestral groups. p.R1628P frequency was highest within the mixed-ancestry group (11.0%, 12/109) (7 of the 11 patients reported having some Chinese ancestry), followed by Chinese (8.3%, 161/1930), Malay (7.7%, 39/507) and Indigenous (3.9%, 4/103) patients.

**Figure 1.**
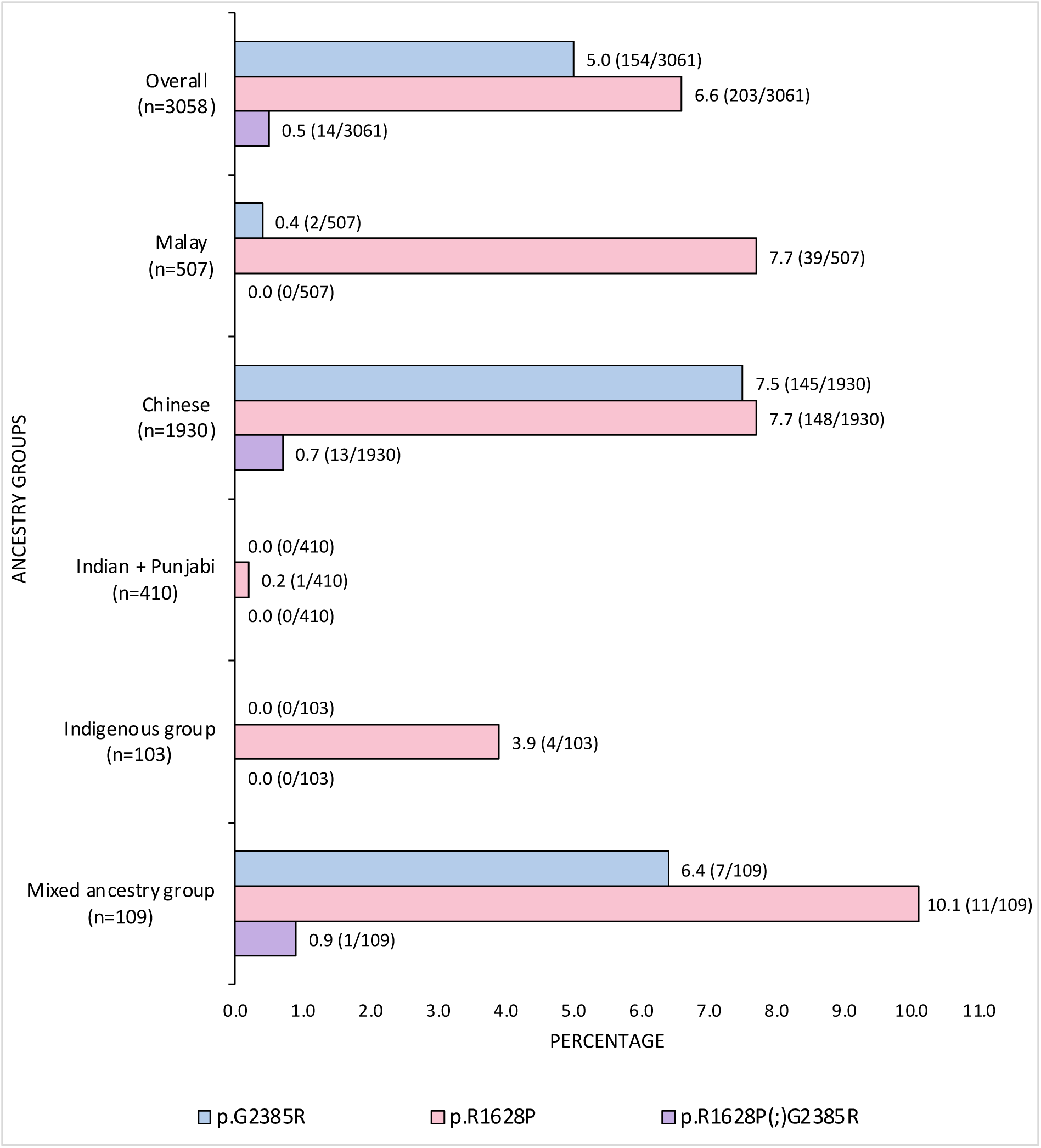
Frequency of *LRRK2* p.G2385R and p.R1628P among PD patients across diverse ancestral groups in Malaysia (n=3,058 index cases). All PD patients who were found to have monogenic PD gene like *PINK1, DJ1, PRKN, SNCA, GCH1*, and *MAPT* were excluded from the analysis. The indigenous groups included ancestral groups such as Bajau, Bidayuh, Bisaya, Bugis, Dusun, Iban, Iranun, Jawa, Kadazan, Kedayan, Lundayoh, Melanau, Murut, Rawa, Rungus, Sinau, Sungai, Suluk, and Tidung.

The mean age at diagnosis of double heterozygous p.R1628P(;)p.G2385R variant carriers (54.4 <u>±</u> 9.1 years) was significantly earlier compared to non-*LRRK2* variant carriers (60.8 <u>±</u> 11.2 years, p=0.031) and p.G2385R carriers (61.7 <u>±</u> 11.1 years, p=0.017), but with no significant difference compared to p.R1628P carriers (59.9 <u>±</u> 10.8 years, p=0.064) (Table 1). There were no significant differences in the mean age at diagnosis between non-carriers, p.R1628P carriers, and p.G2385R carriers. The proportions of EOPD patients (age of diagnosis ≤50) were not significantly different across the four groups.

**Table 1.** Clinico-demographics of PD patients with and without *LRRK2* p.G2385R and p.R1628P in overall cohort (n=3,058).

|  | Non-*LRRK2*-PD<sup>1</sup><br>(n=2,687) | *LRRK2*-PD |  |  | p-value |  |  |  |
| --- | --- | --- | --- | --- | --- | --- | --- | --- |
|  |  | p.G2385R<br>(n=154) | p.R1628P<br>(n=203) | p.R1628P(;)G2385R<br>(n=14) | p.G2385R<br>vs.<br>Non-*LRRK2*-<br>PD | p.R1628P<br>vs.<br>Non-*LRRK2*-<br>PD | p.R1628P<br>(;)G2385R<br>vs.<br>Non-*LRRK2*-<br>PD | p.G2385R<br>vs.<br>p.R1628P |
| **Gender**<br>Male (%) | 58.2 | 53.2 | 56.7 | 64.3 | 0.221<sup>b</sup> | 0.657<sup>b</sup> | 0.648<sup>b</sup> | 0.522<sup>b</sup> |
| **Age at**<br>**Diagnosis**<br>(years) | 60.8 ± 11.2 | 61.7 ± 11.1 | 59.9 ± 10.8 | 54.4 ± 9.1 | 0.332<sup>a</sup> | 0.259<sup>a</sup> | 0.031<sup>a*</sup> | 0.119<sup>a</sup> |
| **EOPD<sup>2</sup>** (%) | 15.8 | 13.1 | 19.6 | 28.6 | 0.359<sup>b</sup> | 0.165<sup>b</sup> | 0.259<sup>c</sup> | 0.104<sup>b</sup> |
| **Presence of**<br>**FH (%)** | 18.7 | 19.4 | 21.1 | 50.0 | 0.821<sup>b</sup> | 0.401<sup>b</sup> | 0.008<sup>c*</sup> | 0.703<sup>b</sup> |

All PD patients who were found to have monogenic PD genes were excluded from the analysis. For families with multiple members affected by PD, only the index cases were included. Normally distributed data were shown as the mean ± standard deviation. Statistical analysis of data was indicated by specific letters: <sup>a</sup>denotes interpretation by T-test, <sup>b</sup>by Chi-Square test and <sup>c</sup>by Fisher's Exact test. Significant differences between groups were denoted by an asterisk (\*). Abbreviations used in the table include: *LRRK2* (*Leucine-Rich Repeat Kinase 2*), PD (Parkinson's Disease), EOPD (Early Onset Parkinson's Disease), FH (Family History).
<sup>1</sup>Non-*LRRK2*-PD was used here to refer to PD patients who were genotyped and found to be negative for both *LRRK2* p.G2385R and p.R1628P risk variants.
<sup>2</sup>EOPD was used to refer to PD patients who were diagnosed before the age of 50 years.

Double heterozygous p.R1628P(;)p.G2385R variant carriers (50%, 7/14) were more likely to have a family history of PD compared to non-carriers (18.7%, 470/2515, p=0.008), p.R1628P carriers (21.1%, 41/194, p=0.021), and p.G2385R carriers (19.4%, 28/144, p=0.016).

### Cross-sectional comparison of motor and non-motor features

Deep clinical motor and non-motor phenotyping was performed in patients recruited for the MJFF Asian *LRRK2* study, comprising a subset of 185 PD patients (n=61 p.R1628P carriers, n=57 p.G2385R carriers, n=5 double heterozygous p.R1628P(;)p.G2385R variant carriers, n=62 non-carriers) (see Methods - Clinical assessments). Their clinico-demographic characteristics are summarized in Table 2. There were no significant between-group differences in age, gender, disease duration, comorbidities (Supplementary Figure 1) or treatment profile (Supplementary Figure 2).

**Table 2.** Detailed phenotyping of motor and non-motor features in PD patients with and without *LRRK2* p.G2385R and p.R1628P (n=185).

|  | Non-*LRRK2*-PD<sup>1</sup> | *LRRK2*-PD |  |  | p-value |  |  |  |
| --- | --- | --- | --- | --- | --- | --- | --- | --- |
|  |  | p.G2385R<br>(n=57) | p.R1628P<br>(n=61) | p.R1628P(;)G2385R<br>(n=5) | p.G2385R<br>vs.<br>Non-*LRRK2*-PD | p.R1628P<br>vs.<br>Non-*LRRK2*-PD | p.R1628P<br>(;)G2385R<br>vs.<br>Non-*LRRK2*-PD | p.G2385R<br>vs.<br>p.R1628P |
| **Age** (years) | 70.3 ± 9.2 | 69.5 ± 9.2 | 67.5 ± 8.5 | 70.4 ± 8.4 | 0.674<sup>a</sup> | 0.090<sup>a</sup> | 0.974<sup>a</sup> | 0.218<sup>a</sup> |
| **Gender**<br>Male (%) | 53.2 | 49.1 | 49.2 | 80.0 | 0.655<sup>c</sup> | 0.654<sup>c</sup> | 0.370<sup>d</sup> | 0.995<sup>c</sup> |
| **BMI** (kg/m<sup>2</sup>) | 23.8 ± 5.7 | 23.3 ± 4.6 | 23.7 ± 4.6 | 25.2 ± 10.2 | 0.571<sup>a</sup> | 0.908<sup>a</sup> | 0.626<sup>a</sup> | 0.613<sup>a</sup> |
| **Presence of FH**<br>(%) | 17.7 | 24.6 | 21.3 | 80.0 | 0.439<sup>d</sup> | 0.626<sup>d</sup> | **0.002<sup>d\*</sup>** | 0.179<sup>d</sup> |
| **Age of Diagnosis**<br>(years) | 61.9 ± 9.2 | 61.9 ± 9.8 | 60.0 ± 9.0 | 58.0 ± 7.3 | 0.988<sup>a</sup> | 0.249<sup>a</sup> | 0.361<sup>a</sup> | 0.280<sup>a</sup> |
| **Disease Duration**<br>(years) | 8.0 [10.0] | 6.0 [8.0] | 6.0 [8.0] | 9.0 [17.0] | 0.343<sup>b</sup> | 0.257<sup>b</sup> | 0.747<sup>b</sup> | 0.823<sup>b</sup> |
| **Constipation Severity**<br>BSC (1-7)<br>PAC-SYM (0-48) | 3.0 [2.0]<br>5.0 [9.0] | 4.0 [1.0]<br>5.0 [8.0] | 4.0 [1.0]<br>5.0 [5.8] | 2.0 [3.0]<br>6.0 [12.5] | 0.118<sup>b</sup><br>0.493<sup>b</sup> | 0.101<sup>b</sup><br>0.777<sup>b</sup> | 0.348<sup>b</sup><br>0.913<sup>b</sup> | 0.780<sup>b</sup><br>0.680<sup>b</sup> |
| **MoCA**<br>Total (0-30)<br>Score<26 (%) | 25.0 [7.0]<br>52.5 | 24.5 [8.0]<br>53.6 | 25.0 [7.0]<br>48.3 | 24.0 [5.0]<br>75.0 | 0.820<sup>b</sup><br>0.904<sup>c</sup> | 0.224<sup>b</sup><br>0.650<sup>c</sup> | 0.967<sup>b</sup><br>0.618<sup>d</sup> | 0.412<sup>b</sup><br>0.573<sup>c</sup> |
| **MDS-UPDRS<sup>4</sup>**<br>Part 1 (0-52)<br>Part 2 (0-52)<br>Part 3 (0-132)<br>Part 4 (0-24)<br>Total (0-260) | 7.5 [11.0]<br>12.5 [16.0]<br>39.0 [25.0]<br>2.0 [6.0]<br>63.5 [44.0] | 8.0 [10.0]<br>9.0 [10.0]<br>43.5 [19.0]<br>2.0 [5.0]<br>66.5 [27.0] | 9.0 [8.0]<br>11.0 [14.0]<br>40.0 [20.0]<br>1.0 [6.0]<br>60.0 [39.0] | 7.0 [15.0]<br>17.0 [26.0]<br>54.5 [27.0]<br>1.5 [7.0]<br>93.0 [85.0] | 0.636<sup>b</sup><br>0.151<sup>b</sup><br>0.112<sup>b</sup><br>0.702<sup>b</sup><br>0.696<sup>b</sup> | 0.339<sup>b</sup><br>0.216<sup>b</sup><br>0.696<sup>b</sup><br>0.667<sup>b</sup><br>0.703<sup>b</sup> | 1.000<sup>b</sup><br>0.358<sup>b</sup><br>0.063<sup>b</sup><br>0.877<sup>b</sup><br>0.138<sup>b</sup> | 0.731<sup>b</sup><br>0.636<sup>b</sup><br>0.129<sup>b</sup><br>0.992<sup>b</sup><br>0.473<sup>b</sup> |
| **Hoehn & Yahr** | 2.0 [1.0] | 3.0 [1.0] | 2.0 [1.0] | 3.0 [2.0] | 0.257<sup>b</sup> | 0.982<sup>b</sup> | 0.093<sup>b</sup> | 0.244<sup>b</sup> |
| **CISI-PD<sup>4</sup>**<br>Motor signs (0-6) | 3.0 [2.0] | 3.0 [1.0] | 3.0 [2.0] | 4.5 [2.0] | 0.529<sup>b</sup> | 0.957<sup>b</sup> | 0.067<sup>b</sup> | 0.576<sup>b</sup> |
| MRC (0-6) | 1.0 [3.0] | 1.0 [2.0] | 1.0 [2.0] | 2.0 [2.5] | 0.381<sup>b</sup> | 0.393<sup>b</sup> | 0.725<sup>b</sup> | 0.976<sup>b</sup> |
| Disability (0-6) | 3.0 [1.0] | 3.0 [1.0] | 3.0 [1.0] | 4.0 [2.5] | 0.466<sup>b</sup> | 0.880<sup>b</sup> | 0.138<sup>b</sup> | 0.536<sup>b</sup> |
| Cognition (0-6) | 2.0 [1.0] | 2.0 [1.0] | 2.0 [2.0] | 2.5 [2.5] | 0.805<sup>b</sup> | 0.453<sup>b</sup> | 0.078<sup>b</sup> | 0.338<sup>b</sup> |
| Total (0-24) | 9.0 [6.0] | 9.0 [4.8] | 10.0 [4.5] | 11.5 [5.5] | 0.885<sup>b</sup> | 0.738<sup>b</sup> | 0.078<sup>b</sup> | 0.658<sup>b</sup> |
| **Phenotype<sup>4</sup> (%)** |  |  |  |  |  |  |  |  |
| Tremor Dominant | 25.0 | 29.1 | 18.6 | 0.0 | 0.582<sup>c</sup> | 0.275<sup>c</sup> | 0.511<sup>d</sup> | 0.389<sup>c</sup> |
| PIGD | 68.3 | 60.0 | 66.1 | 100.0 |  |  |  |  |
| Intermediate | 6.7 | 10.9 | 15.3 | 0.0 |  |  |  |  |
| **LEDD<sup>4</sup> (mg/day)** | 425.3 [525.0] | 457.8 [431.9] | 450.0 [424.5] | 659.3 [389.3] | 0.771<sup>b</sup> | 0.779<sup>b</sup> | 0.462<sup>b</sup> | 0.949<sup>b</sup> |

Normally distributed data were shown as the mean ± standard deviation, while non-normally distributed data were represented by median [interquartile range]. Statistical analysis of data was indicated by specific letters: <sup>a</sup>denotes interpretation by t-test, <sup>b</sup>by Mann-Whitney test, <sup>c</sup>by Chi-Square test and <sup>d</sup>by Fisher's Exact test. Significant differences between groups were denoted by an asterisk (\*). Abbreviations used in the table include: *LRRK2* (*Leucine-Rich Repeat Kinase 2*), PD (Parkinson's Disease), BMI (Body Mass Index), FH (Family History), PIGD (Postural Instability Gait Difficulty), BSC (Bristol Stool Chart), PAC-SYM (Patient Assessment of Constipation-Symptoms), MoCA (Montreal Cognitive Assessment), MDS-UPDRS (The International Parkinson and Movement Disorder Society-Unified Parkinson's Disease Rating Scale), CISI-PD (Clinical Impression of Severity Index for Parkinson's Disease), LEDD (Levodopa Equivalent Daily Dose), and MRC (Motor Response Complications).
<sup>1</sup>Non-*LRRK2*-PD was used here to refer to PD patients who were genotyped and found to be negative for both *LRRK2* risk variants (p.G2385R, p.R1628P).
<sup>2</sup>Other ancestral groups include Melanau, mixed ancestral group, Siamese, and Sikh.
<sup>3</sup>The PD motor phenotype was determined by dividing the mean of MDS-UPDRS items 2.10, 3.15a, 3.15b, 3.16a, 3.16b, 3.17a, 3.17b, 3.17c, 3.17d, 3.17e, and 3.18 by the mean of MDS-UPDRS items 2.12, 2.13, 3.10, 3.11, and 3.12. A ratio greater than 1.15 indicates a tremor dominant phenotype, while a ratio less than 0.90 indicates a PIGD phenotype (Stebbins et al. Mov Disord 2013)
<sup>4</sup>Patients on device aided therapies (DBS, apo-Morphine) were excluded in the analysis.

The frequency of motor features did not differ significantly across the four genotypic groups (Figure 2). There were also no differences in MDS-UPDRS parts II and III scores, H&Y staging, and CISI-PD scores (Table 2). Similar results were observed when all three *LRRK2* Asian risk variant groups were combined into one *LRRK2* group and comparing with non-carriers (data not shown), with no significant differences in motor features, MDS-UPDRS parts II and III scores, H&Y staging, CISI-PD scores, and motor-predominance phenotype. There were no significant between-group differences in the prevalence of motor response complications (Figure 2) and MDS-UPDRS Part IV scores (Table 2). There was also no significant difference in the prevalence of motor response complications between *LRRK2* variant carriers combined (69.1%, 85/123) vs. non-carriers (61.3%, 38/62, p=0.288).

**Figure 2.**
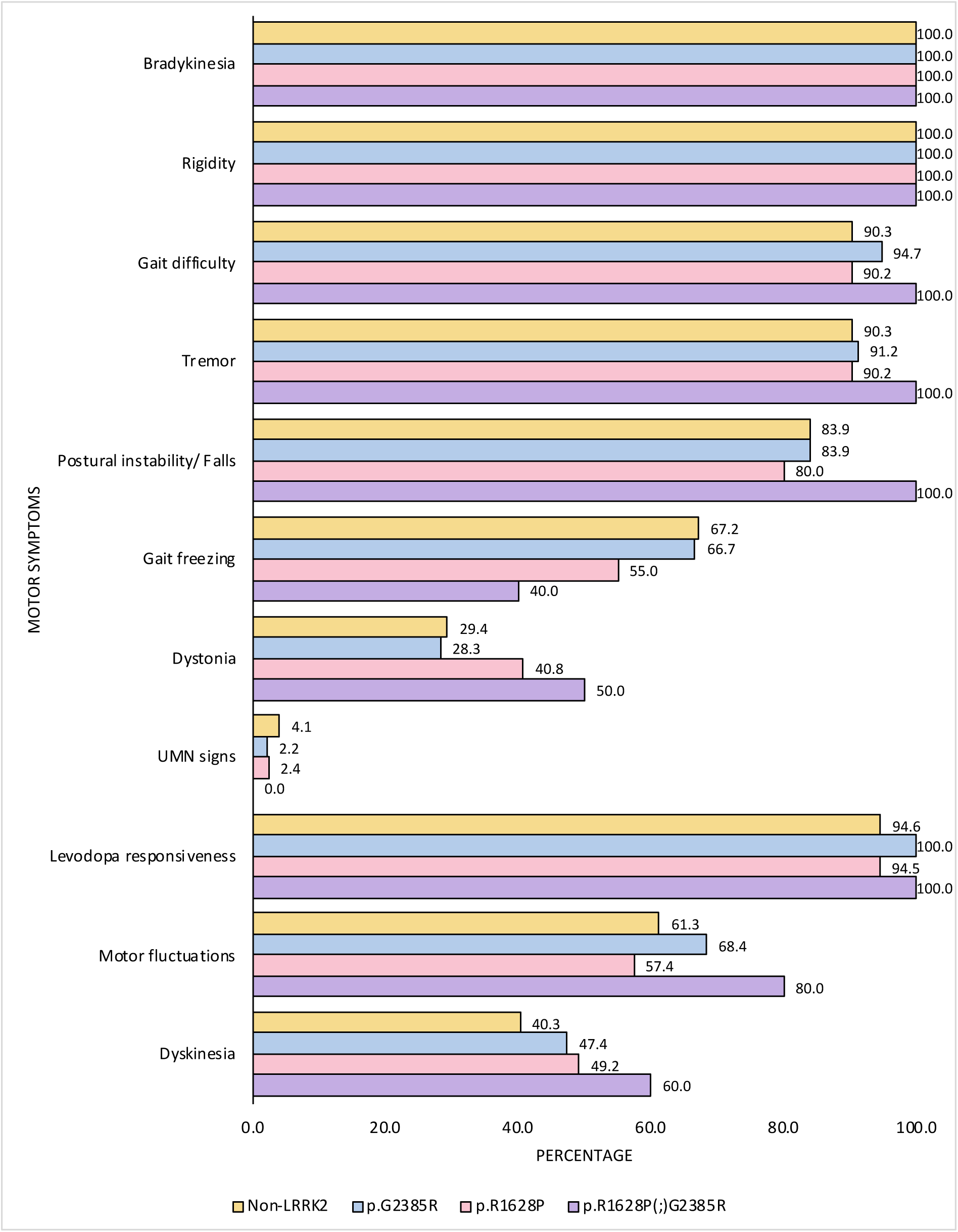
Frequency of motor symptoms and motor response complications in PD patients with and without *LRRK2* p.G2385R and p.R1628P (n=185). Upper motor neuron signs indicate presence of either clonus, hyperreflexia, or abnormal plantar response.

The frequency of most non-motor features (i.e., pain, REM sleep behavior disorder (RBD) symptoms, constipation, mild cognitive impairment (MCI), urinary dysfunction, insomnia, excessive daytime sleepiness, visual hallucinations, hyposmia, anxiety, depression, underweight and impulsive-compulsive behaviors) as well as non-motor symptom burden (MDS UPDRS part 1) did not differ between groups (Figure 3, Table 2). Notably, p.R1628P carriers had a significantly lower prevalence of dementia (2.3%, 1/43) compared to non-carriers (16.3%, 8/49, p=0.034). Dementia frequency in p.G2385R carriers (6.8%, 3/44, p=0.206) and double heterozygous variant carriers (33.3%, 1/3, p=0.442) was not significantly different vs. non-carriers. When combining the p.R1628P, p.G2385R, and double heterozygous p.R1628P(;)p.G2385R variant carriers into a single group, variant carriers appeared less likely to have dementia (5.6%, 5/90) compared to non-carriers (16.3%, 8/49), although the difference did not reach statistical significance (p=0.063). Meanwhile, there were no significant differences in median MoCA scores between variant carriers vs. non-carriers.

**Figure 3.**
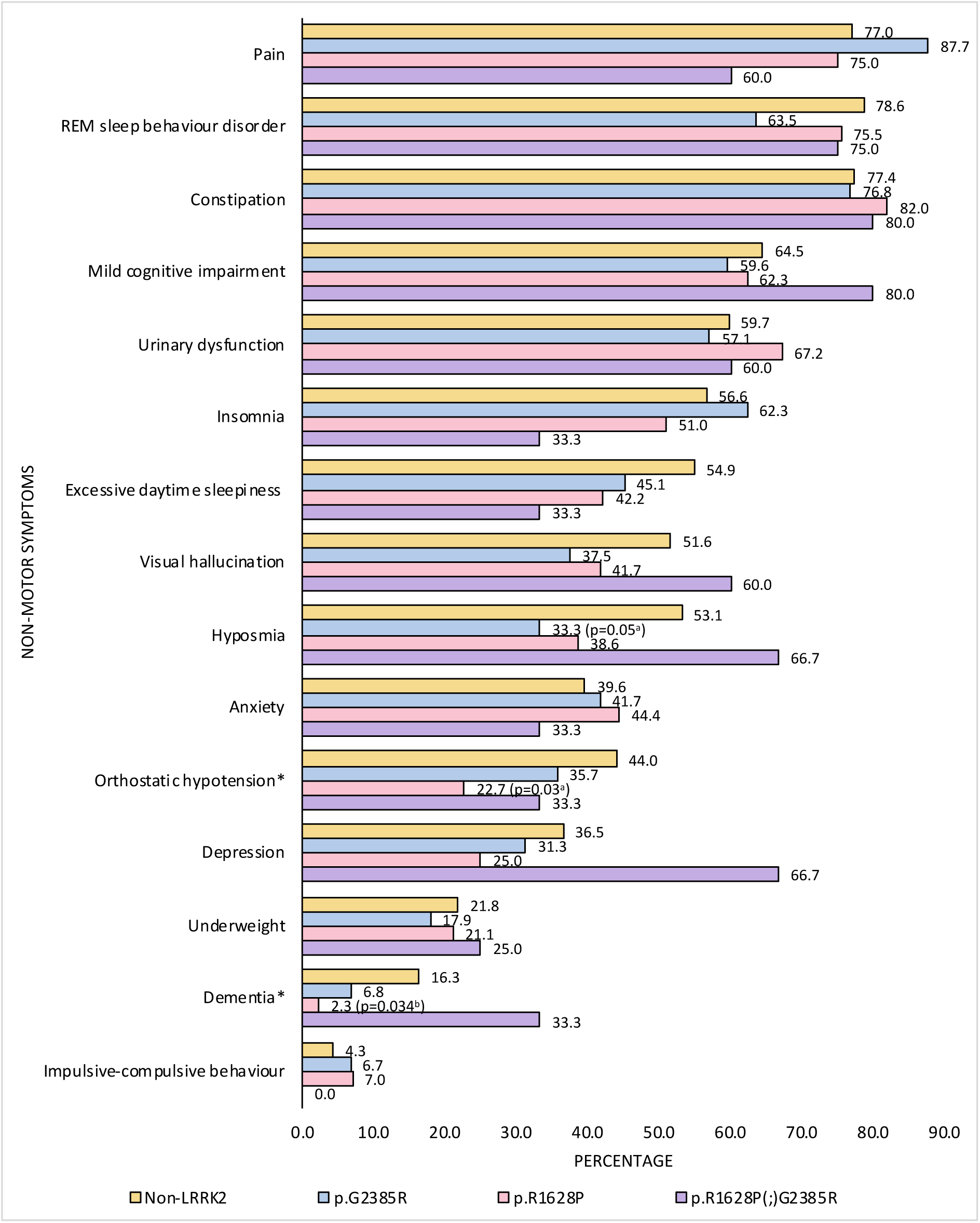
Frequency of non-motor symptoms in PD patients with and without *LRRK2* p.G2385R and p.R1628P (n=185). Significant differences between groups were denoted by an asterisk (*).

Additionally, p.R1628P carriers had a significantly lower frequency of orthostatic hypotension (22.7%, 10/44) compared to non-carriers (44.0%, 22/50, p=0.030). Orthostatic hypotension frequency in p.G2385R carriers (35.7%, 15/42, p=0.523) and double heterozygous p.R1628P(;)p.G2385R variant carriers (33.3%, 1/3, p=1.000) was not significantly different vs. non-carriers. When combining p.R1628P, p.G2385R, and double heterozygous p.R1628P(;)p.G2385R variant carriers into a single group, there were no significant difference in frequency of orthostatic hypotension in variant carriers (29.2%, 26/89) compared to non-carriers (44.0%, 22/50, p=0.078).

### Longitudinal comparison of clinical progression

A total of 635 patients, comprising 539 (84.9%) non-carriers, 47 (7.4%) p.R1628P carriers, 44 (6.9%) p.G2385R carriers, and 5 (0.8%) double heterozygous variant carriers, were followed over a median of 39.0 [23.0] (range:18-65) months using CISI-PD scores to assess clinical disease progression (Table 3). The mean age at the time of baseline CISI-PD assessment was 65.0 ± 9.9 (range:31-89) years.

**Table 3.** Clinical progression of PD patients with and without LRRK2 p.G2385R and p.R1628P (n=635).

|  | Non-*LRRK2*-PD<sup>1</sup><br>(n=539) | *LRRK2*-PD |  |  | p-value |  |  |  |
| --- | --- | --- | --- | --- | --- | --- | --- | --- |
|  |  | p.G2385R<br>(n=44) | p.R1628P<br>(n=47) | p.R1628P(;)G2385R<br>(n=5) | p.G2385R<br>vs.<br>Non-*LRRK2*-PD | p.R1628P<br>vs.<br>Non-*LRRK2*-PD | p.R1628P<br>(;)G2385R<br>vs.<br>Non- *LRRK2*-PD | p.G2385R<br>vs.<br>p.R1628P |
| **Age** (years) | 64.9 ± 10.0 | 66.5 ± 10.5 | 63.9 ± 9.1 | 67.2 ± 4.6 | 0.344<sup>a</sup> | 0.453<sup>a</sup> | 0.622<sup>a</sup> | 0.205<sup>a</sup> |
| **Gender**<br>Male (%) | 54.5 | 50.0 | 57.4 | 60.0 | 0.561<sup>c</sup> | 0.702<sup>c</sup> | 1.000<sup>d</sup> | 0.476<sup>c</sup> |
| **Presence of FH**<br>(%) | 24.9 | 27.9 | 28.3 | 80.0 | 0.657<sup>c</sup> | 0.610<sup>c</sup> | **0.016<sup>d\*</sup>** | 0.970<sup>c</sup> |
| **Age of Diagnosis**<br>(years) | 61.7 ± 10.6 | 63.7 ± 10.5 | 61.1 ± 9.2 | 60.6 ± 7.8 | 0.288<sup>a</sup> | 0.716<sup>a</sup> | 0.822<sup>a</sup> | 0.216<sup>a</sup> |
| **Disease Duration**<br>(years) | 2.0 [5.0] | 1.0 [4.0] | 2.0 [4.0] | 4.0 [12.0] | 0.840<sup>b</sup> | 0.573<sup>b</sup> | 0.655<sup>b</sup> | 0.869<sup>b</sup> |
| **Mean Follow-Up Duration**<br>(months) | 39.0 [22.0] | 38.5 [26.0] | 34.0 [22.0] | 44.0 [25.0] | 0.521<sup>b</sup> | 0.105<sup>b</sup> | 0.458<sup>b</sup> | 0.605<sup>b</sup> |
| **CISI-PD Baseline** |  |  |  |  |  |  |  |  |
| Motor signs (0-6) | 3.0 [1.0] | 3.0 [2.0] | 3.0 [2.0] | 4.0 [2.0] | 0.504<sup>b</sup> | 0.861<sup>b</sup> | **0.021<sup>b\*</sup>** | 0.699<sup>b</sup> |
| MRC (0-6) | 0.0 [2.0] | 0.0 [2.0] | 0.0 [2.0] | 1.0 [3.0] | **0.019<sup>b\*</sup>** | 0.573<sup>b</sup> | 0.337<sup>b</sup> | 0.165<sup>b</sup> |
| Disability (0-6) | 3.0 [0.0] | 3.0 [0.0] | 3.0 [1.0] | 4.0 [3.0] | 0.500<sup>b</sup> | 0.229<sup>b</sup> | **0.028<sup>b\*</sup>** | 0.139<sup>b</sup> |
| Cognition (0-6) | 1.0 [1.0] | 1.0 [2.0] | 1.0 [1.0] | 2.0 [3.0] | 0.975<sup>b</sup> | 0.904<sup>b</sup> | 0.099<sup>b</sup> | 0.953<sup>b</sup> |
| Total (0-24) | 8.0 [4.0] | 7.0 [4.0] | 8.0 [4.0] | 12.0 [5.0] | 0.491<sup>b</sup> | 0.573<sup>b</sup> | **0.015<sup>b\*</sup>** | 0.923<sup>b</sup> |
| **CISI-PD Motor**<br>(prog/month) | 0.021 [0.037] | 0.009 [0.037] | 0.000 [0.026] | 0.019 [0.024] | 0.294<sup>b</sup> | **0.012<sup>b\*</sup>** | 0.436<sup>b</sup> | 0.266<sup>b</sup> |
| **CISI-PD MRC**<br>(prog/month) | 0.000 [0.034] | 0.000 [0.046] | 0.000 [0.036] | 0.000 [0.168] | 0.725<sup>b</sup> | 0.429<sup>b</sup> | 0.846<sup>b</sup> | 0.426<sup>b</sup> |
| **CISI-PD Disability**<br>(prog/month) | 0.019 [0.029] | 0.000 [0.020] | 0.007 [0.024] | 0.000 [0.048] | **0.001<sup>b\*</sup>** | **0.005<sup>b\*</sup>** | 0.244<sup>b</sup> | 0.886<sup>b</sup> |
| **CISI-PD Cognition**<br>(prog/month) | 0.021 [0.042] | 0.021 [0.047] | 0.000 [0.037] | 0.029 [0.038] | 0.951<sup>b</sup> | 0.252<sup>b</sup> | 0.993<sup>b</sup> | 0.366<sup>b</sup> |
| **CISI-PD Total**<br>(prog/month) | 0.068 [0.077] | 0.048 [0.094] | 0.050 [0.087] | 0.058 [0.139] | 0.264<sup>b</sup> | **0.011<sup>b\*</sup>** | 0.816<sup>b</sup> | 0.422<sup>b</sup> |

Normally distributed data were shown as the mean ± standard deviation, while non-normally distributed data were represented by median [interquartile range]. Statistical analysis of data was indicated by specific letters: <sup>a</sup>denotes interpretation by t-test, <sup>b</sup>by Mann-Whitney test, <sup>c</sup>by Chi-Square test and <sup>d</sup>by Fisher's Exact test. Significant differences between groups were denoted by an asterisk (\*). Abbreviations used in the table include: *LRRK2* (*Leucine-Rich Repeat Kinase 2*), PD (Parkinson's Disease), and CISI-PD (Clinical Impression of Severity Index for Parkinson's Disease).
<sup>1</sup>Non-*LRRK2* PD was used here to refer to PD patients who were genotyped and found to be negative for both *LRRK2* risk variants (p.R1628P, p.G2385R).
<sup>2</sup>Other ancestral groups include Sinokadazan, Suluk, and mixed ancestral groups.
<sup>3</sup>The CISI-PD progression rates were determined by calculating the difference between the latest and the baseline CISI-PD scores, then dividing this difference by the follow-up period in months and were reported as change in scores per month (i.e., prog/month). Patients on device aided therapies (DBS, apo-Morphine) were excluded in the analyses of CISI-PD.

The p.R1628P carriers had a slower progression of total CISI-PD scores compared to non-carriers (increase of 0.050 points per month [0.087] vs. 0.068 [0.077], p=0.011). The progression of total CISI-PD scores was not significantly different in p.G2385R carriers (0.048 [0.094], p=0.264) and double heterozygous p.R1628P(;)p.G2385R variant carriers (0.058 [0.139], p=0.816) vs. non-carriers.

When analyzing component domains of the CISI-PD, the p.R1628P carriers exhibited slower progression in motor signs compared to non-carriers (0.000 [0.026] vs. 0.021 [0.037], p=0.012), as well as in disability (0.007 [0.024] vs. 0.019 [0.029], p=0.005). The p.G2385R carriers also exhibited slower progression in disability (0.000 [0.020], p=0.001) vs. non-carriers.

When the *LRRK2* variant carriers were combined, there was a slower progression in the total CISI-PD score (0.049 [0.092] vs. 0.067 [0.077], p=0.013), as well as in motor signs (0.000 [0.028] vs. 0.021 [0.037], p=0.011), and disability (0.000 [0.020] vs. 0.019 [0.029], p<0.001).

After controlling for baseline age and corresponding CISI-PD scores (i.e., when analyzing total CISI-PD, this was adjusted for baseline total CISI-PD score, and when analyzing the motor signs component score, in turn this was adjusted for baseline motor signs score, and so on), differences between the p.R1628P carriers vs. non-carriers in total CISI-PD (p=0.009), motor signs (p=0.014), and disability (p=0.001) progression scores remained significant, as did the difference between p.G2385R carriers vs. non-carriers in the disability progression scores (p=0.001).

## DISCUSSION

In this large multicenter Malaysia-wide study involving more than 3000 multiethnic Asian PD patients, we found a high frequency of the two *LRRK2* Asian risk variants, p.G2385R and p.R1628P, with 12.1% harboring at least one variant. We confirmed a relatively high frequency (8.2%) of the *LRRK2* p.G2385R variant in patients of Chinese ancestry. In addition, while it is less studied, the p.R1628P variant was in fact overall more common than p.G2385R in our cohort, with Chinese having a frequency of 8.3% and Malays having a frequency of 7.7%. The presence of the p.R1628P variant is also now reported for the first time in Malaysian indigenous patients, which was found in a considerable proportion (3.9%) of patients. Notably, these variants were very rare (0.2%, only one p.R1628P carrier) among Indian patients. Overall, *LRRK2* variant carriers, particularly those with p.R1628P, appeared to have a more “benign” disease course, with a lower prevalence of significant non-motor features (dementia and orthostatic hypotension) that are known to have a substantial impact on quality of life and survival in PD patients.^60–62^ They also had lesser longitudinal changes in global clinical disease severity, particularly in the domains of motor signs and disability over a median observation period of ∼40 months. Double heterozygous p.R1628P(;)p.G2385R variant carriers had significantly earlier age at disease onset, on average in their mid-50s, compared to single variant or non-carrier groups who were typically diagnosed in the late 50s or early 60s.

The overall prevalence of 5.0% of *LRRK2* p.G2385R carriers and 6.6% of *LRRK2* p.R1628P carriers in the present study are consistent with findings from our previous Malaysian study a decade ago, but now involving a much larger (>4-fold) sample size.^16^ In that study, 5.2% of PD patients were p.G2385R carriers and 6.2% were p.R1628P carriers.^16^ This stability of frequency over time is in keeping with the notion that genetic factors are a relatively stable component contributing to PD pathogenicity (although note that all the patients from the earlier study were also included in the current report).^2,4,63^ The present study highlights significant ethnicity-specific distributions of *LRRK*2 variants among Malaysian PD patients. In Chinese patients, the frequencies of p.G2385R (7.5%) and p.R1628P (7.7%) are comparable to the findings of previous studies, with p.G2385R having an overall prevalence of 10.3% (n=5184) and p.R1628P having an overall prevalence of 5.3% (n=4492) among patients in China,^64^ and very similar to early findings in the Chinese PD population in Singapore (7.5% with p.G2385R, 8.4% with p.R1628P).^17,19^

Among Malay PD patients, we observed a 7.6% frequency for p.R1628P, higher than previously reported in Singapore (2.3%).^11^ Our estimate is likely to be more precise, given the substantially larger sample size (>3-fold; n=500 vs. 132) of Malay patients in the present study. This has important implications, as Malays comprise ∼200 million individuals living in Southeast Asia, spread across Indonesia, Malaysia, Singapore, Brunei, and Southern Thailand, countries that are projected to experience substantial growth in the number of PD cases over the coming decades.^3,54,65^ Interestingly, these Asian *LRRK2* risk variants are almost non-existent among Indians,^26,66^ who are the other major racial group in Malaysia. In the largest pan-India PD genetics study to date (n=4806 patients), p.G2385R and p.R1628P were each found in only 0.1% of patients.^66^ The stark contrast further highlights the importance of recognizing ethno-geographic differences in the underlying etiology, frequency, and clinical presentation of PD and related neurodegenerative or movement disorders.^3,67–69^

We report for the first time the presence of p.R1628P in Malaysian indigenous groups, with an overall frequency of 3.8% among the 103 indigenous PD patients screened. The variant was identified among the Bajau (1/16), Bugis (1/7), Sungai (1/5), and Dusun (1/28) subgroups. The limited number of indigenous patients underscores the need for augmented efforts to study these populations, which have rarely been included in genetics research.^5,47^ Our preliminary and ongoing work in this area has revealed that these populations also harbor pathogenic variants (in *PRKN*, *SNCA*, *PINK1*, etc.) causing monogenic PD and are thus relevant to obtaining a more complete and equitable understanding of PD genetics globally.^53,70^

Regarding demographic features, our findings revealed no significant differences between carriers of single *LRRK2* Asian risk variants and non-carriers in terms of age of diagnosis, consistent with most published studies.^21,29,31,32,38,40–42^ In contrast, we found that double heterozygous p.R1628P(;)p.G2385R variant carriers exhibited a significantly earlier mean age of diagnosis (54.4 ± 9.1 years), which supports the hypothesis that interactions between multiple *LRRK2* risk variants modulate PD pathogenesis.^71^ To our knowledge, only one study, led by researchers in Singapore,^72^ investigated the effect of carrying multiple *LRRK2* Asian risk variants and likewise demonstrated a lower age of PD onset (52.6 ± 12.3 years, vs. 62.5 ± 10.5 years in those without risk variants). This study included p.S1647T in their analysis, but the absence of a significant impact when this variant was combined with either p.G2385R or p.R1628P suggests that the effect may be potentially specific to the p.G2385R-p.R1628P combination. While the frequency of the double heterozygous p.R1628P(;)G2385R variants is relatively low (14/3058, 0.5%), further research into risk variant interactions may yield important biological insights into *LRRK2*-PD pathogenesis.^73^

In addition, our study explored the clinical manifestations of PD in relation to *LRRK2* Asian risk variant carrier status. Although a few studies have reported higher prevalences of PIGD phenotype and motor fluctuations among p.G2385R^35,36^ and *LRRK2* variant carriers,^34–36^ respectively, we did not find significant differences in motor symptoms and motor response complications between carriers and non-carriers, which aligns with the bulk of the published literature^21,29,30,33,38,41^. Our findings also accord with the largest study to date profiling clinical features in Asian *LRRK2* variant carriers (Song et al. studied not only p.G2385R [n=304] and p.R1628P [n=220], but also p.A419V [n=105]), which found no overall motoric (UPDRS II and III) differences in *LRRK2* variant carriers vs. idiopathic PD. The exception was that in those with p.G2385R, tremor was slightly milder at baseline (by about half a UPDRS point, presumably using a composite score for both rest and action tremor, in which case the range of scores would be 0-28). We previously reported that the application of DBS treatment (which is primarily aimed at treating motor response complications, and to some extent also tremor) did not seem to be over-represented among carriers of *LRRK2* risk variant carriers, in contrast to *GBA1* mutation carriers.^74^

While there is limited literature investigating the non-motor features of PD among *LRRK2* risk variant carriers,^18,21,32,34,36,41–43^ we found that p.R1628P carriers exhibited a lower frequency of dementia and orthostatic hypotension compared to non-carriers. To our knowledge, these are novel observations and require confirmation in other cohorts. Research specifically addressing cognitive impairment among Asian *LRRK2* risk variant carriers remain very limited,^18,21,32,34,36,41,43^ with two studies reporting higher MMSE scores among p.G2385R carriers,^34,36^ with another reporting a higher frequency of “cognitive impairment”, based on MMSE scores, among a very small cohort (n=11) of p.R1628P carriers.^41^ Other studies of monogenic *LRRK2*, involving p.G2019S and p.R1441C, reported a lower prevalence of dementia compared to non-carriers.^8,75,76^ A possible explanation for these observations is that cognitive dysfunction is associated with the burden of Lewy body pathology,^77,78^ in PD in general as well as in *LRRK2*-PD specifically,^79^ and *LRRK2* mutation carriers are often negative for synucleinopathy. For example, Siderowf and colleagues recently reported that ∼1/3 of PD patients carrying p.G2019S were negative on the cerebrospinal fluid synuclein seed amplification (SAA) test.^49,80^ Intriguingly, one small neuropathological study reported that there was significantly less highly-aggregated α-synuclein in the brains of patients with p.G2019S compared with idiopathic PD.^81^ Similarly, although no previous study has looked into the prevalence of orthostatic hypotension among p.R1628P carriers, studies of other *LRRK2* variants (involving p.G2019S and p.R1441G) have shown that autonomic dysfunction, including orthostatic hypotension, which is thought to reflect more widespread synucleinopathy affecting the nervous system,^82^ is less common than in idiopathic PD.^8,83^ Interestingly, researchers have also documented a mechanistic link between orthostatic hypotension and cognitive impairment and dementia in PD, probably related to repeated cerebral hypoperfusion.^61,84,85^ The study by Song et al.^37^ reported that in their p.G2385R carrier group, fewer patients had excessive daytime sleepiness (EDS) at baseline compared to idiopathic PD (28.6% vs. 36.6%). In our study, EDS was less frequently reported in variant carriers (45.1%, 42.2%, and 33.3% in p.G2385R, R1628P, and p.R1628P(;)G2385R, respectively, vs. 54.9% in non-carriers), but the differences were not statistically significant. Standardized use of the Epworth Sleepiness Scale may have enhanced the ability of Song and colleagues to detect this difference.^86^ In PD, a relationship between EDS and cognitive impairment, as well as generally in advancing disease, have long been recognized.^87–89^

Previous studies on disease progression in p.G2385R and p.R1628P carriers were often limited by sample size, with 10-21 variant carriers in each group;^44,45^ one study combined the pathogenic p.G2019S and p.R1441C/G variants with p.R1628P under a general *LRRK2* group in their analyses.^46^ These methodological limitations make interpretation of the data challenging.^44–46^ The largest study to date, by Song et al.^37^, did not find differences in overall motor (UPDRS II and III) progression, either with the *LRRK2* variants analyzed separately or when combined, but tremor progressed less in the p.G2385R carriers, over a mean follow-up of 5.6 (± 1.3) years. Our study found that *LRRK2* variant carriers exhibited less global disease progression over time, particularly in terms of motor signs (p.R1628P) and disability (p.G2385R and p.R1628P). The differences in study findings may be due to several factors, including a substantially longer mean disease duration at baseline in previous studies (5-6 years,^37,44^ vs. median 2 years in our study), as well as their longer follow-up duration (4-6 years,^37,44,45^ vs. slightly >3 years in ours). Furthermore, different assessment tools were utilized. Our study employed the CISI-PD, while prior research used the MDS-UPDRS, UPDRS, and Hoehn and Yahr scales,^37,44^ although the literature suggests that these scales are highly correlated.^90^ Overall, however, when giving greater weightage to the larger studies, a picture seems to be emerging of a slightly more “benign” progression of disease with the *LRRK2* Asian variants compared with idiopathic PD, akin to what has been observed with p.G2019S.^7,8^

A major strength of this work is that it is by far one of the largest studies on *LRRK2*-related PD in Southeast Asia, encompassing a diverse multiethnic cohort that includes Chinese, Malay, Indian, and indigenous populations. It is also the first to report on *LRRK2* variants in indigenous Malaysian groups, contributing novel insights into the genetic epidemiology of PD in these very underrepresented populations.^5,47^ Furthermore, our study is one of a few which comprehensively investigated a broad range of non-motor features, using a standardized checklist,^9,54,56,57^ among *LRRK2* Asian variant carriers compared to non-carriers. Additionally, we were able to include a large number of *LRRK2* variant carriers and non-carrier patients (∼100 and >500, respectively) in the longitudinal analysis of PD progression. Importantly, these patients represented a broad spectrum of disease duration and severity and provided, we believe, a “real-world” sampling of PD patients, since the CISI-PD is brief and non-onerous to patients. The scale could thus be applied consecutively to every patient, including those unable (e.g., due to severe disease) to complete more laborious clinical evaluations.^91,92^ To our knowledge, longitudinal progression data for the CISI-PD has only been reported once, from Spain,^93^ and the results presented here (which are comparable to the Spanish data), involving the largest sample size to date with *n*>600, can serve as a valuable “real-world” reference dataset.

Some study limitations need to be acknowledged. We were unable to exclude the possibility that other genetic factors might also play a role. Additionally, the follow-up period for this report of our ongoing study was not very long, which limited the ability to observe longer-term disease progression. The designation of dementia was made in the clinic setting, rather than on a research basis utilizing expert neuropsychological evaluations. Future studies should aim for comprehensive genetic and genomic screening, longer follow-up periods, expanded sample sizes for rarer subgroups, incorporation of technology-based tools such as sensors and advanced analytics that can enhance “deep phenotyping” of patients,^94,95^ and biomarker studies,^10,51,73^ to better delineate the impact of *LRRK2* variants on PD genetic epidemiology, phenotype, and biology.

## CONCLUSION

In conclusion, our findings highlight the high prevalence of two *LRRK2* Asian risk variants in Malaysia and a relatively more “benign” disease course among p.R1628P carriers. Our findings are highly relevant in the era of personalized medicine and can be utilized for better disease prognostication and management. The high frequency of *LRRK2* p.G2385R and p.R1628P variants in Malaysia should be considered when broadening the recruitment of participants for studies of *LRRK2*, since such underrepresented populations with enriched cases of “genetic PD” may also be able to host, and benefit from, genetics-targeted biomarker and therapeutic programs.

## AUTHOR CONTRIBUTIONS

A.H.T. and S.-Y.L. conceived the idea and designed the study. A.H.T., S-Y.L., J.P.S., Y.Y.B., K.A.I., A.S.M., T.T.L., I.L., Y.K.C., J.C.E.O., W.C.L., S.C.W., Y.H.L., P.C.T., T.L.O., W.K.C. contributed to patient recruitment, clinical data, and biological sample collection. J.W.G., R.Y.O., Q.H.Y., C.C.Y.L., T.S.T., J.W.H., Y.W.T., J.Z., A.N.K.A., H.X.D., and L.C.L. contributed to clinical data collection, data and biobanking management. J.L.L., Y.W.T., J.W.G., and A.A-A. contributed to the genotyping and genetic analysis. J.W.G. and A.H.T. conducted the data analyses. J.W.G., S.-Y.L., A.H.T., and A.A-A. performed manuscript writing. All authors reviewed the manuscript for revision and approved the final manuscript.

## CORRESPONDING AUTHORS

Correspondence and request for materials should be addressed to Ai Huey Tan MD PhD FRCP or Shen-Yang Lim MD FRACP or Azlina Ahmad-Annuar PhD.

## ACKNOWLEDGEMENTS

The authors gratefully acknowledge funding from the Michael J. Fox Foundation for Parkinson’s Research (MJFF), which supported a collaborative research effort between the University of Malaya, Malaysia, and the University of Dundee, United Kingdom, for the discovery of Asian *LRRK2* biomarkers (MJFF-010188, MJFF-021041, MJFF-022659, awarded to AHT, SYL, and ES). We would also like to thank Samantha Hutten and Rebecca Ouellette from MJFF for their kind assistance and support in the project and grant management. The authors gratefully acknowledge Mr. and Mrs. Ooi Teong Siew for their kind and generous assistance in purchasing laboratory equipment. The authors gratefully acknowledge the patients and their families for participation in this study.

## DATA AVAILABILITY STATEMENT

The data generated and analyzed in this study, including genetic and clinical phenotype data of Parkinson’s disease patients with the *LRRK2* p.G2385R and p.R1628P risk variants, are available from the corresponding author upon reasonable request. Due to ethical and privacy considerations, direct public access to the dataset is restricted. Any requests for data access should be directed to.

## Declaration

All authors declare no competing interests.

