## Supplementary Figure for "*LRRK2* p.G2385R and p.R1628P Variants in a Multi-Ethnic Asian Parkinson’s Cohort: Epidemiology and Clinical Insights"

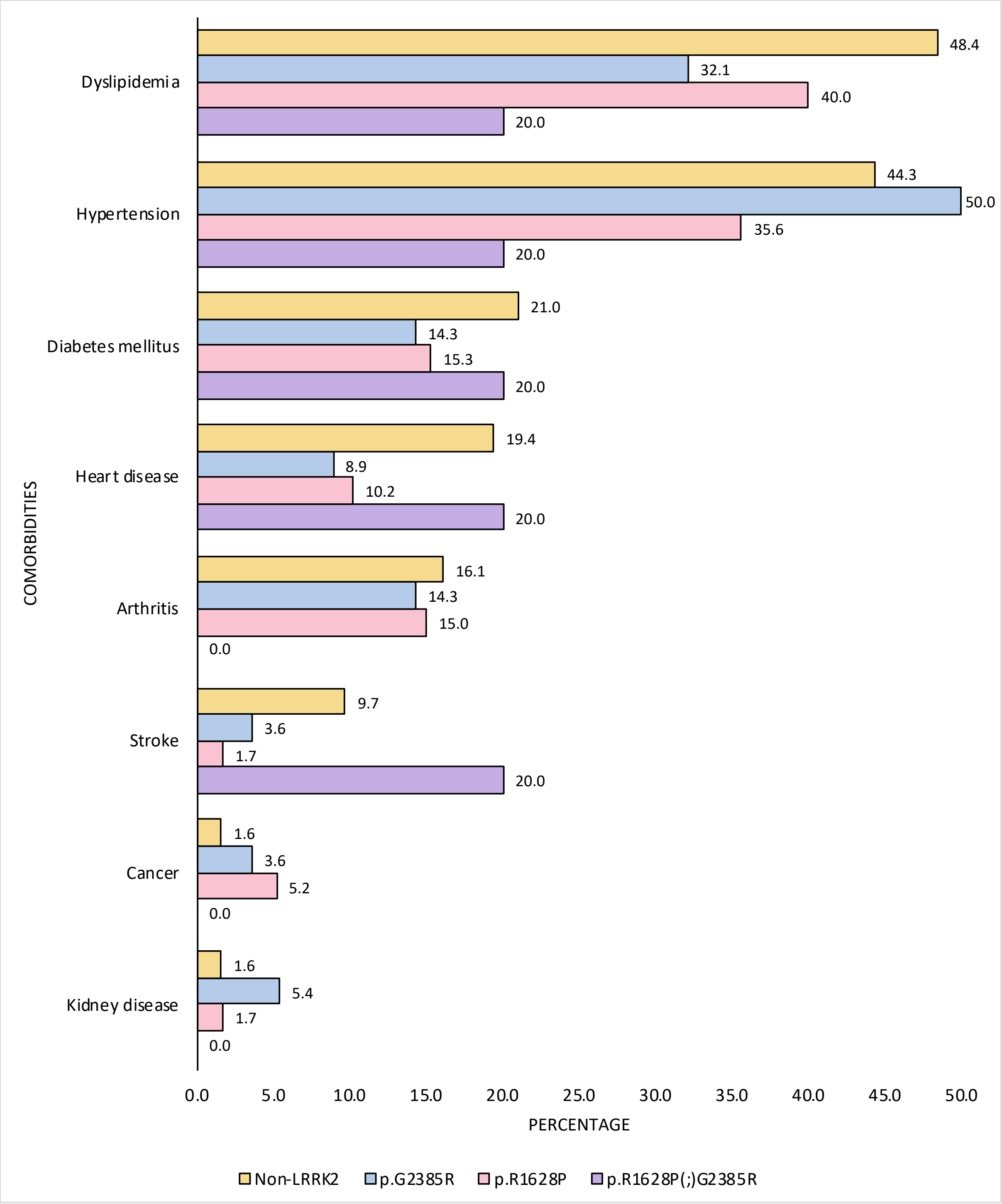


**Supplementary Figure 1. Frequency of comorbidities in PD patients with and without *LRRK2* p.G2385R and p.R1628P.** There was only one *LRRK2* p.R1628P PD patient diagnosed with inflammatory bowel disease.

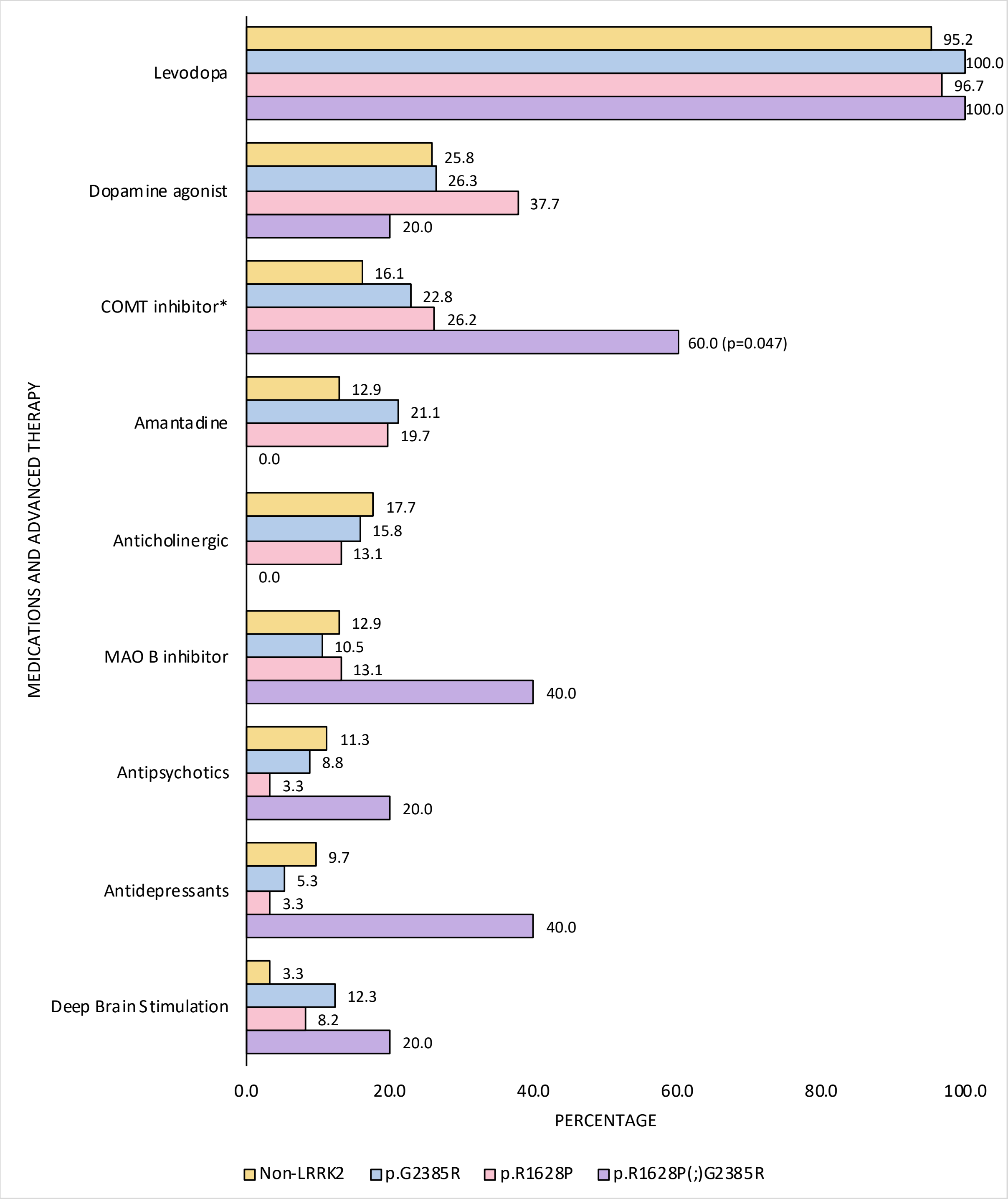


**Supplementary Figure 2. Treatment profile (oral medications and advanced therapies) in PD patients with and without *LRRK2* p.G2385R and p.R1628P (n=185).** There was only one patient (non-*LRRK2*-PD) on apomorphine infusion. Significant differences between groups were denoted by an asterisk (*). Abbreviations used in the figure include: COMT (Catechol-O-methyltransferase), MAO-B (Monoamine Oxidase B).
